# Tracking Inflammation in Real Time Following Vaccination: Validation of a Novel Individualized Digital Inflammatory Biomarker Relative to Serum Biomarkers

**DOI:** 10.1101/2025.10.13.25337893

**Authors:** Darpit Dave, Ryan Heumann, Stephan Wegerich, Jadranka Sekaric, Jaap Oostendorp, Robert Paris, Matthew P. Ward, Steven R. Steinhubl

## Abstract

**Background:** Inflammatory changes underly many diseases and therapeutic interventions, making accurate tracking of inflammation critical for clinical evaluation of disease course and therapy response. Traditional methods like fever detection and serum biomarkers are limited by imprecision and invasiveness. Collection of self-reported symptoms after vaccination is a common vaccine trial endpoint, but prone to bias. Wearable sensors offer a promising alternative by detecting subtle physiological changes over time. Prior studies show they identify transient post-vaccine inflammation but lack validation relative to serum biomarkers.

**Methods:** This study included 61 volunteers who were administered one of four mRNA vaccines (1 or 2 doses) for a total of 80 doses. Participants wore a torso sensor patch for 14 days beginning seven days before vaccination, whose data was used to derive an individualized digital biomarker of inflammation – inflammatory multivariate change index (iMCI). The reference outcome was a serum biomarker panel collected at baseline and five post-vaccination timepoints. Self-reported reactogenicity symptoms were tracked daily for 7 days starting the day of vaccination. The correlation between total iMCI response within 48 hours following vaccination and maximal change in select serum biomarkers was determined, along with their relationship to reactogenicity.

**Findings:** There was a moderate to strong positive Spearman correlation between total iMCI and change in C-reactive protein (CRP) (0.59, p < 0.01) and interferon gamma (IFN-_Y_)(0.56, p < 0.01) across vaccine types and vaccine doses, similar to the correlation between CRP and IFN-_Y_ (0.60, p < 0.01). The associations with self-reported systemic reactogenicity was only moderate for all: 0.48, p < 0.01 for iMCI, 0.34, p = 0.01 for interferon gamma, 0.36, p <. 0.01 for C-reactive protein.

**Interpretation:** The personalized multivariate inflammatory digital biomarker derived from wearable sensor data can quantify an individual’s inflammatory response to vaccination as an alternative to serial serum biomarker testing. This scalable, non-invasive approach can enable real-time monitoring of the onset, duration, and severity of inflammation.

**Funding:** *Moderna, Inc*

## INTRODUCTION

Inflammation due to immune system activation plays a central role in a range of acute and chronic disease processes such as infections, autoimmune conditions, cancers, and atherosclerosis.^1^ Furthermore, the effectiveness of many therapeutic interventions’ effectiveness depends on their ability to either deactivate the immune system and decrease inflammation, such as disease-modifying antirheumatic drugs (DMARDs),^2^ or temporarily activate it and increase inflammation such as immune-oncology therapies^3^ and vaccines.^4^ Therefore, the ability to accurately track changes in an individual’s inflammatory state, especially subtle changes, could play a vitally important role in early disease detection and therapeutic responsiveness.

Development of a fever, an imprecise population-based measure of a robust inflammatory response defined over 150 years ago^5^, is the most common clinical measure of systemic inflammation, but is not useful for detecting mild changes. Beyond that, the only common objective measurement of change in a person’s inflammatory status requires serial measurement of serum inflammatory biomarkers, such as C-reactive protein (CRP), which is invasive, inconvenient and, therefore, assessed infrequently. The innate inflammatory response following vaccination is unique in that the symptoms due to inflammation – termed reactogenicity – are frequently followed, especially during clinical trials.^6^ However, the subjective and self-report nature of reactogenicity makes it susceptible to the nocebo effect, which has been shown to explain over 50% of reported systemic symptoms.^7^

Wearable sensors offer a possible alternative method for objectively measuring small but clinically relevant changes in a person’s inflammatory state. By continuously tracking an individual’s normal variability in their physiology at rest and during activity over multiple days, small deviations from that “normal” following an inflammatory stimulus, such as a vaccine, can be detected. Multiple studies using commercial wearable sensors have confirmed their ability to detect subtle but significant individual changes following vaccination, primarily against COVID-19.^8–10^ However, the majority of these studies were limited by restricting the wearable data analyzed to a daily metric, evaluating the post-vaccine changes in single data types, and few including simultaneous serum inflammatory biomarkers.^11^

We recently reported the performance of a multivariate digital biomarker, developed using data from a torso sensor patch and a machine learning method of similarity-based modeling (SBM), in a real-world, observational study of 88 people who received a mRNA vaccine against COVID-19.^12^ We found that this novel digital biomarker better predicted the presence of systemic reactogenicity than any single data type, and, in a subgroup, correlated with vaccine-induced immunogenicity. Here, we build on these results in a new study of this individualized multivariate digital biomarker in healthy volunteers receiving four different mRNA vaccines in a Phase-1 study and evaluate its relationship to serially collected serum inflammatory biomarkers and reactogenicity symptoms.

## METHODS

### Study design and participants

This study is a sub-study of a larger Phase-1 study of four different mRNA vaccines and FLUAD as an active influenza comparator.^13^ The protocol was approved by the Advarra Institutional Review Board, Columbia, MD and registered at ClinicalTrials.gov: NCT05397223. All participants provided written informed consent for participation in the study.

This study enrolled a total of 61 volunteers with an average age of 50 (range 18 – 73) and 61% female. Participants received either a single 50 µg dose of mRNA-1273 (bivalent coronavirus disease of 2019 – COVID-19), mRNA-1345 (respiratory syncytial virus – RSV), or mRNA-1010 (influenza), or two 100 µg doses of mRNA-1647 (cytomegalovirus CMV), or a standard 0.5 ml dose of FLUAD (active influenza comparator).

Across participants, 80 vaccine doses were administered, but after excluding individuals and doses without analyzable data, 53 participants and 65 unique vaccine doses were available for analysis. Individuals were excluded from analysis due to inadequate serum biomarker data (n=13) or unavailable patch data (n=3). The study population is summarized in **Table 1**. Participants who received FLUAD (n=5 total, 3 with analyzable data) were also excluded from this analysis to maintain consistency with mRNA vaccine-induced inflammation.

**Table 1:** Age, sex and vaccine doses of participants.

| Vaccine Type | Percentage<br>Female (%) | Median Age<br>(Range) | Participants Vaccinated<br>(Participants with<br>Analyzable Data) |  |
| --- | --- | --- | --- | --- |
|  |  |  | Dose 1 | Dose 2 |
| mRNA-1010 (Influenza HA) | 70.0 | 49 (30 – 68) | 10 (10) | 0 |
| mRNA-1273 (SARS-CoV-2) | 58.3 | 57 (32 – 62) | 12 (10) | 0 |
| mRNA-1345 (RSV) | 66.7 | 50 (19 – 73) | 21 (18) | 0 |
| mRNA-1647 (CMV) | 64.0 | 38 (18 – 47) | 14 (12) | 14 (12) |
| FLUAD | 60.0 | 53 (21 – 57) | 5 (3) | 0 |
| **TOTAL** | **61.00** | **50 (18 – 73)** | **66 (53)** | **14 (12)** |

### Serum biomarkers

A total of 63 serum biomarkers were potentially measurable at each of the six time points: baseline (pre-vaccine) blood-draw was conducted 0.5-1 hour before the vaccine dose, and post-vaccine blood draws were collected at 6-12 hours, 24-48 hours, 72-96 hours, one week, and then 4-weeks after vaccine. The 63 biomarkers were narrowed down to six for primary analysis. These serum biomarkers are C-reactive protein (CRP), interferon gamma (IFN-_Y_), Chemokine Ligand 9 (CXCL9), Chemokine Ligand 10 (CXCL10), Macrophage inflammatory protein-1 beta (MIP-1 beta), Monocyte Chemotactic Protein 2 (MCP-2). These biomarkers were selected based on the data quality criteria, including low rates of missingness and presence of clean, numerically interpretable values (excluding ambiguous or non-numeric entries such as ‘<value’, ‘quantity not sufficient (QNS)’, and ‘not reported (NR)’), visual inspection of consistent patterns of change among participants and published evidence of a serum biomarker’s association with vaccineinduced inflammation.^14–17^ **(Supplemental Figure 1)** Out of the six selected serum biomarkers, CRP and IFN-_Y_ were our primary focus due to their previously established role in clinical medicine and vaccine-induced inflammatory response.^18^ Serum biomarker response is defined as the maximum change in individual serum biomarkers levels (at any of the post-vaccine visits) from the baseline value.

### Wearable sensor

All study participants wore a VitalPatch^TM^ by VitalConnect (San Jose, CA) during the course of the study. This sensor is a 510(k)-cleared, wearable, disposable adhesive patch with a battery life of seven days.

The patch collects electrocardiogram (ECG), tri-axial accelerometry, skin temperature, and posture information. These physiology signals are transferred to the native mobile app via a Bluetooth connection which are uploaded and synced with a cloud-based server using the internet. When using cellular networks, a transport layer security (TLS) cryptographic protocol was applied between the mobile phones and the server to ensure a secure transfer. No personal identifiers were stored or transmitted at any point during the data sync or from the mobile app. The cloud-based platform was securely hosted in the Google cloud and analytics server. All data were de-identified, and reference was provided using a participant ID. The data could only be obtained or viewed via secured authenticated login. In addition to the patch sensor, participants also wore a modified Samsung Galaxy smartwatch, but those data are not incorporated into this analysis.

### Individualized Digital Inflammatory Biomarker

The inflammatory multivariate digital biomarker evaluated in this study was developed by combining multiple physiological parameters into a single aggregated metric derived from the patch data, as described in detail in our previous work reference.^12^ At its core, the digital biomarker uses machine learning methodologies within a similarity-based modeling (SBM) framework, which learns the dynamic interplay between the multivariate input sources. The input sources included continuous measurements of skin temperature, heart rate, heart rate variability, respiration rate and activity level, all derived from the biosensor waveform data collected via the patch. Personalized baseline models of each participant’s unique physiological dynamics are established, creating a ‘digital twin’, which when compared to new input data following vaccination, removes expected variations, leaving only inflammation-induced differences, known as residuals. These residuals are generated for each of the physiology data stream and then combined into a single aggregated metric, inflammatory multivariate change index (iMCI), which is normalized to range between 0 and 1, with 1 indicating highest degree of inflammatory change, and updated every 15-minutes.

The totality of a volunteer’s inflammatory response to a vaccine was determined by employing an area under the curve (AUC) approach. The ‘iMCI total response’ was defined as *A*_i_/*A*_T_, where *A*_T_ is the total rectangular area within the time window and *A*_i_ is the area under the curve for iMCI during the window. The metric is defined for a fixed window of time, here 48 hours, starting from the time of administration of a vaccine dose. The derivation of iMCI from multi-modal physiology data and computing the iMCI total response using the AUC-based window approach is described in detail in reference 15.

### Reactogenicity

Reactogenicity was solicited through a self-reported qualitative survey (e-Diary) of the severity of a total of 10 symptoms, six systemic and four local symptoms. eDiary entries were recorded daily with the first entry starting just after vaccination, and then until six days after. A reactogenicity response metric was developed based on the number of symptoms reported, their respective severity (grades 0 through 3), and the total number of days the sign or symptom was reported. Details on the symptoms recorded, severity grading, computation of the reactogenicity response and its distribution among study participants can be found in **Supplemental Figure 2.**

### Statistical analysis

The primary objective of this study was to evaluate the association, on an individual level, between iMCI from a chest-patch data and serum inflammatory biomarker levels changes following all vaccine doses. A secondary objective was to evaluate the correlation between participant reported reactogenicity, iMCI and serum inflammatory biomarkers. To test these objectives, hypothesis testing was performed to evaluate the correlation between total iMCI individual serum biomarker response and self-reported reactogenicity. This was compared to the correlation between the individual serum biomarkers to each other and to reactogenicity. To check for the strength, and significance of these correlations, we used the non-parametric Spearman’s correlation test. Since normality assumptions were not met using the Shapiro-Wilk test, the non-parametric approach of Spearman’s correlation test was preferred.

## RESULTS

### Individual Multiparametric Digital Inflammatory Biomarker (iMCI)

Changes in individual inflammatory state over time following vaccine as determined by their iMCI response are shown in **Figure 1**. Substantial interindividual variability is noticeable across vaccine-type and dose. Greater iMCI responses were seen in individuals likely to have immunological memory, either via a booster dose or prior infection. For example, most volunteers receiving their 2^nd^ dose of mRNA-1647 tended to have a greater inflammatory response via iMCI compared to other single dose vaccine types. In addition, three of the four volunteers who tested seropositive for prior CMV infection preceding their mRNA-1647 first dose, highlighted in red, had a greater response than seronegative participants.

**Figure 1:**
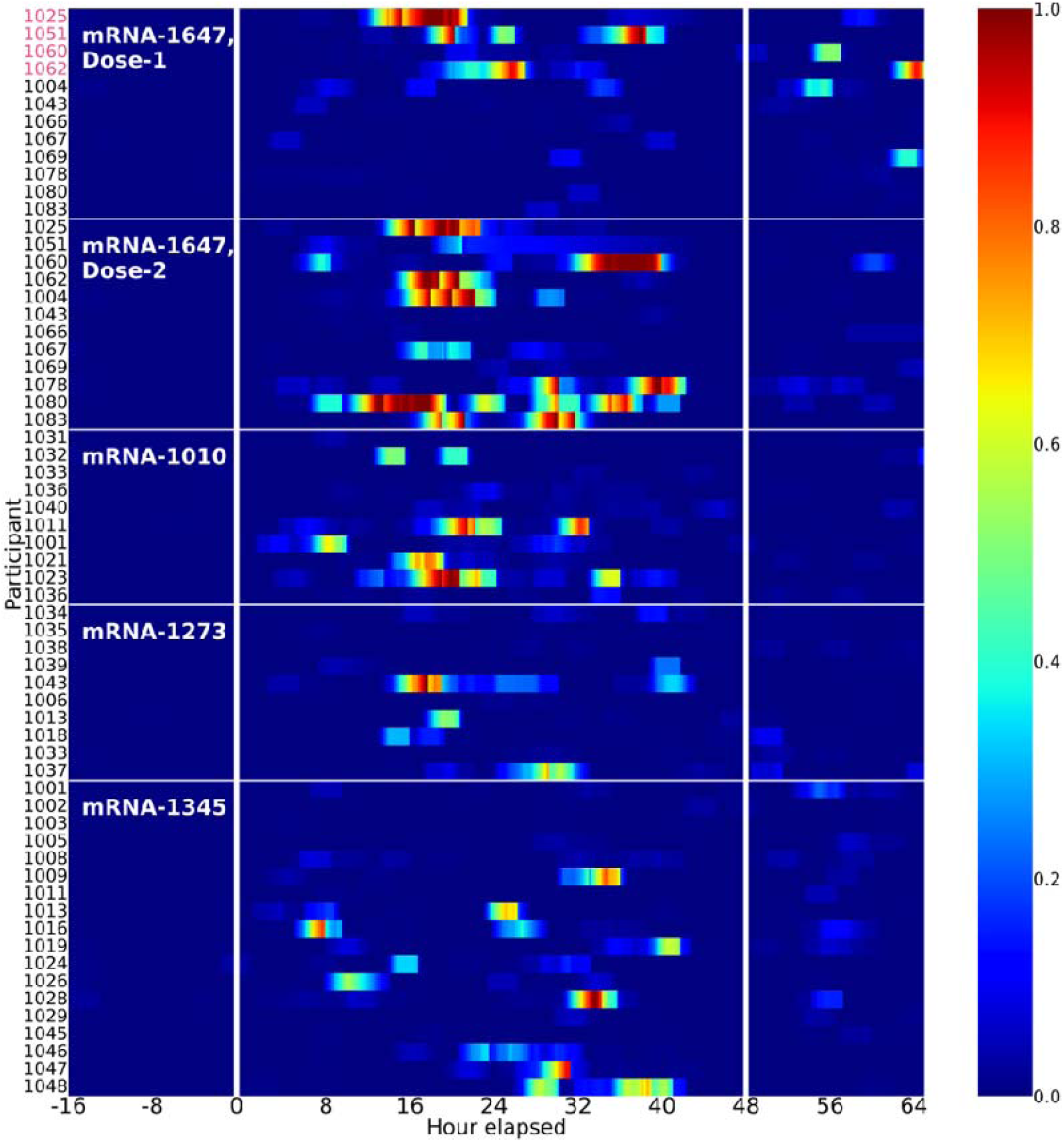
Heatmap for iMCI values for all individuals across vaccine types and doses. Time “0” is the time of vaccination. Vaccines include first and second doses mRNA-1647 against CMV, mRNA-1010 against influenza, mRNA-1273 against COVID-19, and mRNA-1345 against RSV. Participant numbers in red are volunteers who tested sero-positive for CMV prior to vaccination.

The iMCI Total Response was determined for each volunteer as described as a method of summing all the changes shown in the heat map over the 48 hours post-vaccine. As seen in **Figure 2A**, while there was substantial interindividual variability in the iMCI Total Response within each vaccine, individuals receiving a second dose of mRNA-1647 tended to have the greatest response, with equally high levels noted for three of the four CMV seropositive individuals following their first dose of mRNA-1647.

**Figure 2:**
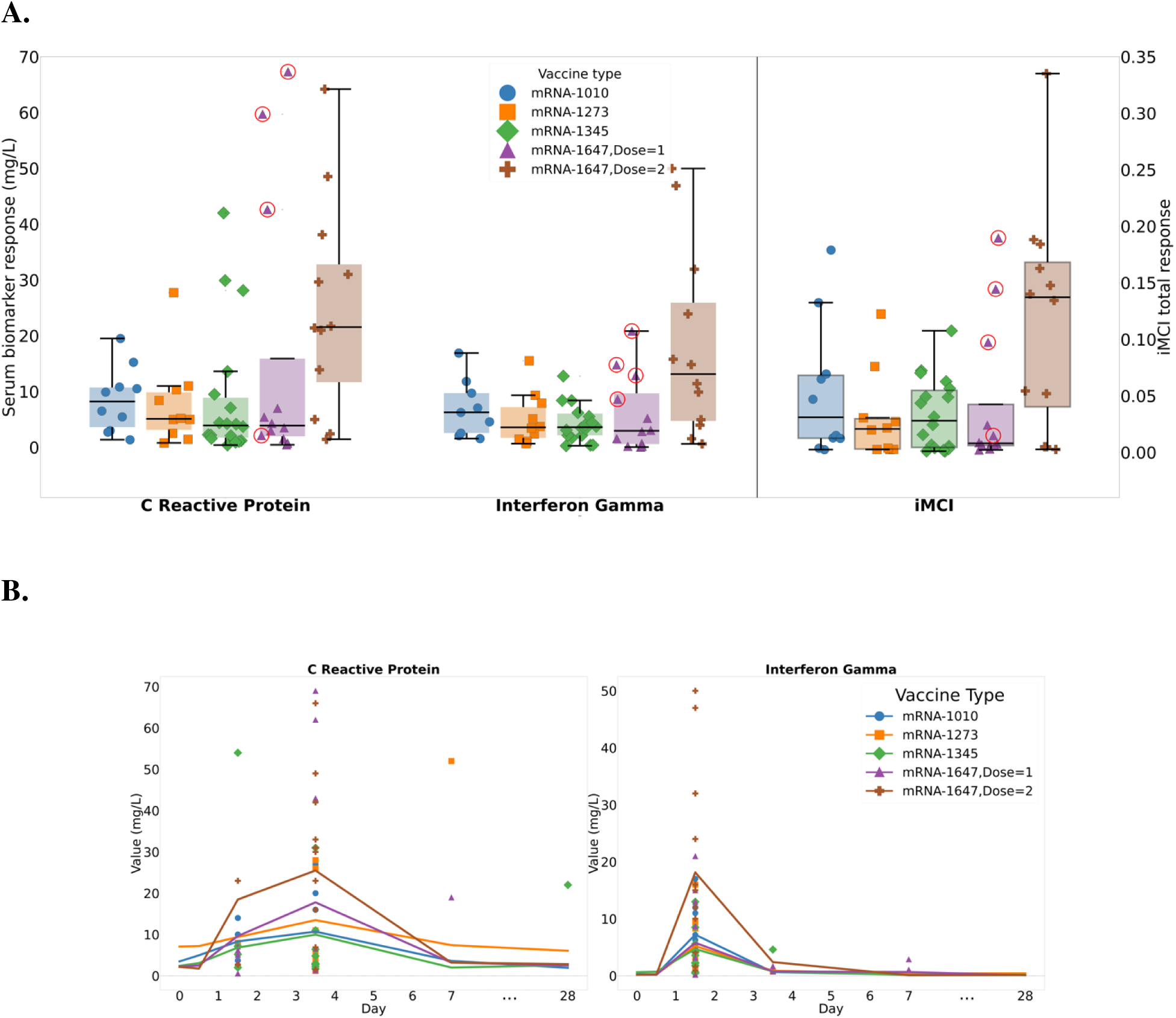
(**A**). Maximal change in CRP, Interferon Gamma and iMCI values across vaccine types/dose. Individuals who were sero-positive for CMV at baseline are circled in red. Boxplots show the overall distribution of the response with the line representing the median value for each vaccine type and dose. Overlayed scatter plots show responses for participants who received a particular vaccine type and dose (**B).** Serum biomarker trajectories for C Reactive Protein and Interferon Gamma across all vaccine types and doses over time. Solid lines represent the mean trajectories for each vaccine type whereas the markers represent the maximal change from baseline for each serum-biomarker response recorded for each individual for that vaccine type and dose.

### Serum biomarkers

For the serum biomarkers, CRP and IFN-_Y_ showed consistent trajectories for all participants across vaccine types and doses. **(Figure 2B)** For IFN-_Y,_ maximal changes from baseline were most commonly seen between 24 to 48 hours following vaccination with return to baseline by day 4. For CRP, peak changes tended to be later, between 72 and 96 hours, with return to baseline by day 7.

Similar to the distribution of iMCI total response, substantial interindividual variability is evident in both CRP and IFN-_Y_ across each vaccine type and dose, with those receiving a second dose of mRNA-1647, and with CMV seropositive participants prior to their first dose tending to have greater responses. **(Figure 2A)**

Consistent trends across the four other serum biomarkers with peak values occurring between the 2^nd^ or 3^rd^ day and serum biomarker levels returning back to pre-vaccine levels between 4-7 days post vaccination **(Supplemental Figure 3).**

### Correlations Between iMCI and Serum Biomarkers

As shown in **Figure 3A**, in pooling all vaccine responses, a moderate to strong correlation was found between iMCI Total Response and the maximal change in CRP and IFN-_Y_ following vaccination. These correlations were statistically significant and monotonically increasing in nature. The strength of the correlation between the digital biomarker and the two serum biomarkers is similar to the strength of the correlation between CRP and IFN-_Y_.

**Figure 3:**
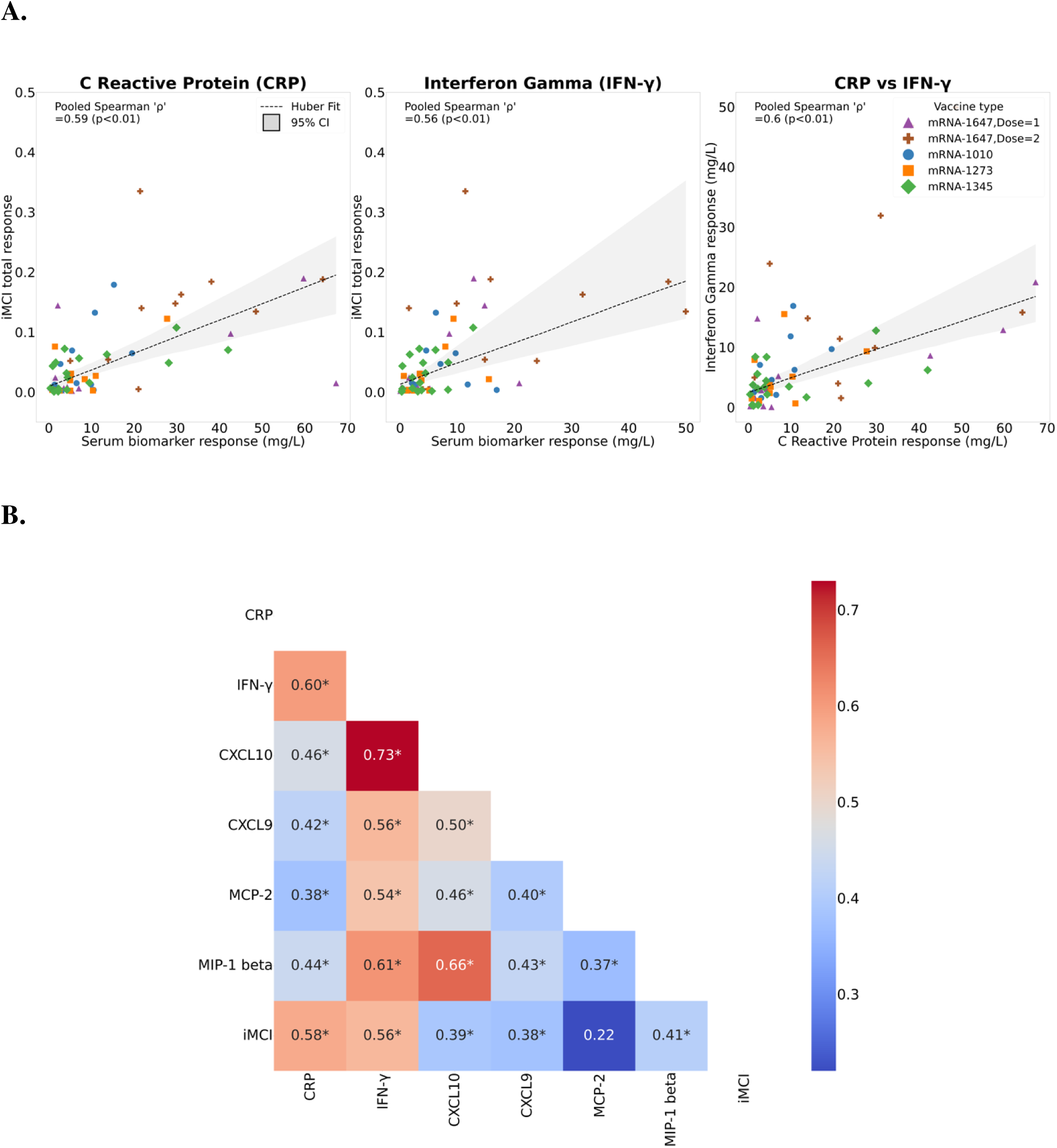
(**A**). Scatterplots of individuals’ iMCI vs IFN-_γ_, iMCI vs CRP, and CRP vs IFN-_γ_ across different vaccine types and doses. Linear relationship was estimated using Huber regression (dashed line), with the shaded region representing the 95% confidence interval based on bootstrapping (**B).** Correlation plot for all mRNA vaccine combined with all six selected serum biomarkers and iMCI Total Response. Numbers in squares indicate the pooled Spearman rho, and an * indicates statistical significance at p<0.05.

iMCI Total Response also had a statistically significant association with all other studied serum biomarkers except Monocyte Chemotactic Protein 2 (MCP-2), however these associations were only moderately strong **(Figure 3B).**

### Reactogenicity

Self-reported systemic reactogenicity responses for all vaccine types and doses are shown in **Figure 4A**. Individual variation across vaccine types and dose are seen. Unlike for serum biomarkers and iMCI total response, individuals receiving dose 2 of mRNA-1647 did not report higher levels of systemic reactogenicity symptoms relative to single-dose vaccines. When evaluating association between serum biomarkers and iMCI to systemic reactogenicity, we found only a moderately strong association, although statistically significant. **(Figure 4B)**

**Figure 4:**
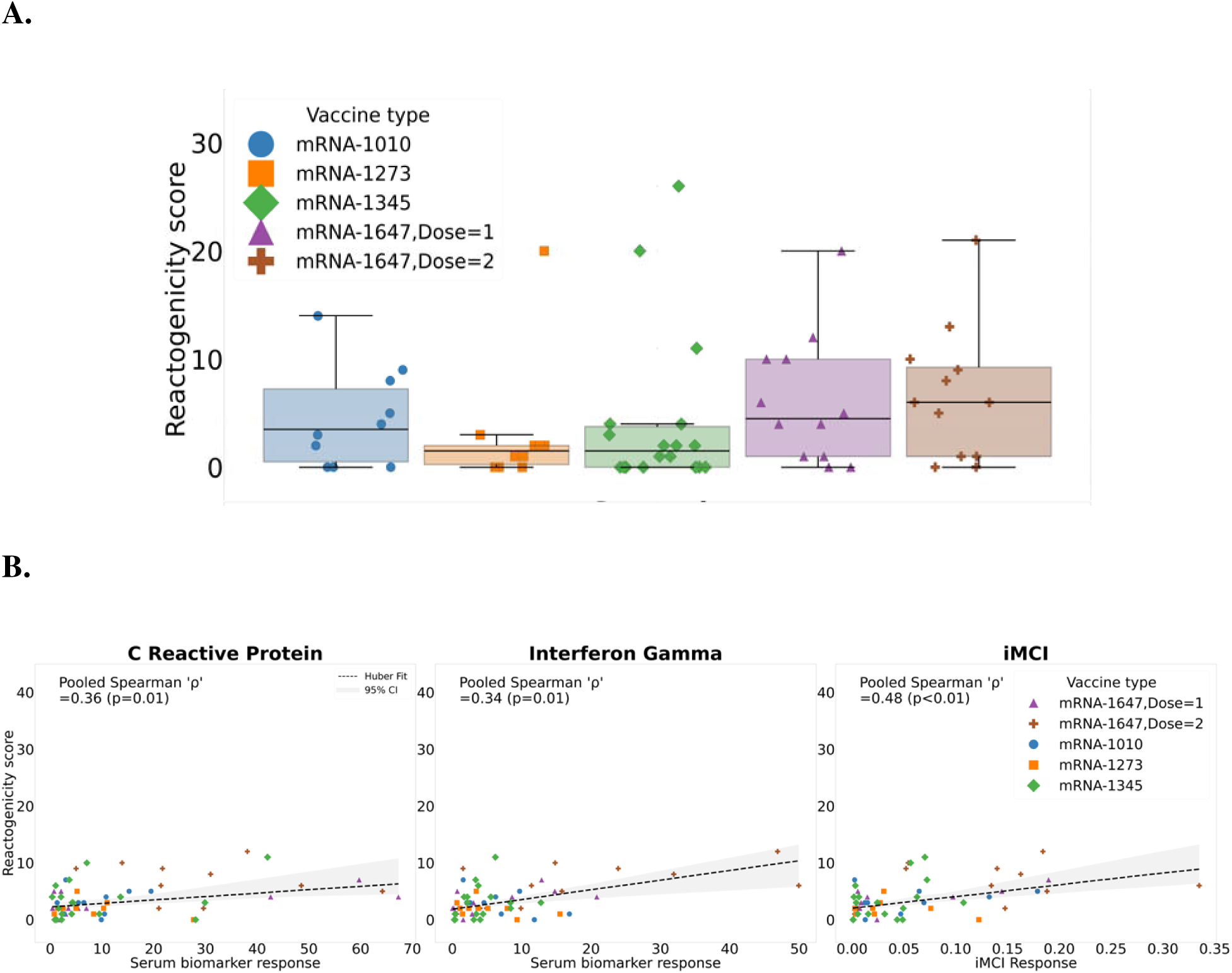
(**A**) Individual systemic reactogenicity scores across vaccine types and dose (**B**) Scatter plots of iMCI total response, maximal change in CRP, and maximal change in IFN-_γ_ versus individual systemic reactogenicity score. The relationship was modeled using a Huber regression (dashed line) with 95% CI derived via bootstrapping (shaded area).

The relationship between iMCI and serum biomarkers with local reactogenicity and total (local + systemic) reactogenicity are shown in **Supplemental Figure 4.**

## DISCUSSION

Wearable sensors are opening new frontiers in healthcare by enabling insights that were previously unattainable. By comparing an individual’s current physiological state to their own unique baseline, these devices can support more precise and personalized diagnostics and care. One particularly promising application is the detection of changes in inflammatory status, given the central role of immune activation and inflammation in health, disease progression, and therapeutic response.^19^

This study is the first to validate the correlation between a multivariate, individualized digital inflammatory biomarker – iMCI – and serum inflammatory biomarkers following innate immune system activation initiated by multiple mRNA vaccine types and doses. It also builds on previous findings that linked this digital biomarker to self-reported reactogenicity following real-world mRNA COVID-19 vaccination.^12^ In the current study, iMCI performed at least as well as serum biomarkers in predicting subjective reactogenicity, supporting that individual physiologic responses to inflammation or infection as detectable through wearable sensors can precede or parallel symptom perception and may offer a more objective and temporally resolved measure.^20^

Our results demonstrate that the iMCI effectively captures the physiological manifestations of vaccine-induced inflammation. Its performance was comparable to that of tracking changes in traditional serum inflammatory biomarkers – particularly CRP and IFN-_Y_ – measured serially after vaccination, suggesting that iMCI may serve as a reliable, non-invasive surrogate for conventional inflammatory assays.^21–23^. Notably, the strength of the correlation between iMCI and these biomarkers was comparable to the correlation observed between the biomarkers themselves.

A wide range of wearable devices exist, each with its own advantages and limitations, but able to detect subtle physiologic changes when normalized to that individual. For example, a recent prospective study incorporated 3 different sensors – a ring, a wrist wearable, and a multi-sensor shirt – and serum inflammatory biomarker tracking to demonstrate how data from different wearables can be used to train machine learning algorithms to predict systemic inflammation following exposure to a live attenuated influenza vaccine.^11^ For our study, we selected a medical-grade patch sensor to ensure the collection of high-fidelity, beat-to-beat physiological data. This data was processed to produce 17 distinct source signals, sampled every minute. We then applied a machine learning technique known as similarity-based modeling to integrate all data streams and their interactions, enabling the creation of a personalized ‘digital twin’ for each participant.^24^ By analyzing these signals in 15-minute intervals, we could objectively track each individual’s unique physiological responses to inflammation. This approach allowed us to continuously compare real-time physiological data against each participant’s baseline model, effectively filtering out expected variations and isolating vaccine-induced changes.

Immune system function is highly variable between people, but relatively stable over time for an individual.^25^ Inflammation, a hallmark of innate immune response, manifests through systemic physiologic changes such as elevated heart rate, increased respiratory rate, reduced heart rate variability (HRV), and elevated skin temperature—all of which can be captured by high-fidelity wearable devices.^26^ Prior work has found that the degree of change in each individual biometric following vaccine-induced inflammation differs widely between people, making the tracking of a single physiologic parameter less sensitive.^12^ The iMCI leverages multiple signals to create a composite, individualized index that reflects the magnitude and dynamics of a person’s inflammatory response.

This study examined inflammation triggered by several mRNA vaccines, highlighting one of several potential applications for a digital inflammatory biomarker. Currently, vaccination strategies follow a “one-size-fits-all” model, wherein nearly all individuals receive the same approved dose of a given vaccine, despite well-documented variability in immune responses.^27^ Emerging evidence suggests that the magnitude and nature of vaccine-induced inflammation may serve as a predictor of immunogenicity.^28^ Therefore, monitoring post-vaccination inflammation could inform dosing strategies to optimize efficacy and safety. Beyond vaccination, a digital biomarker such as iMCI may also support individualized treatment approaches in other immunomodulatory therapies, including chimeric antigen receptor (CAR) T cell therapies and biologic interventions for autoimmune diseases.^29,30^

### Limitations

Several limitations should be considered in interpreting these results. First, the timing of serum biomarker blood draws may have led to missed peak levels, potentially underestimating the true magnitude of the inflammatory response. Second, the sample size was a limited convenience sample, requiring duplication in a larger, more diverse population to confirm and extend these findings. In addition, it should be kept in mind that iMCI is an early prototype. While promising, it can be improved upon with additional refinement and validation. Finally, these results are limited to data available from a patch sensor, which is less usable for longer term monitoring.

Further work using data from a wrist-wearable sensor can potentially expand the implementation of an individual digital inflammatory biomarker once validated.

## Conclusions

Wearable sensors coupled with advanced data analytics offer a novel and scalable approach to quantifying an individual’s inflammatory response to vaccination. In this study we have validated that an individualized digital inflammatory biomarker, iMCI, correlates with serum inflammatory biomarkers following vaccination.

## Data Availability

Requests for de identified summary data from this study can be made by submitting a written request with an analysis plan to the corresponding author for review. Only de identified individual level data may be shared to protect participant privacy, and contingent on IRB approval.

