## Supplementary for "Tracking Inflammation in Real Time Following Vaccination: Validation of a Novel Individualized Digital Inflammatory Biomarker Relative to Serum Biomarkers"

### Reactogenicity

The four local symptoms include injection at the pain site, measurement of injection site erythema, measurements of swelling (hardness) at the injection site, and auxiliary (underarm) swelling or tenderness. The six systemic symptoms include headache, fatigue, myalgia (muscle aches), joint aches, nausea/vomiting, and chills.

Based on the grading scheme, total reactogenicity score is calculated by aggregating across all symptoms and for all days of the study period. For the purpose of our analysis, we consider total reactogenicity (combining local and systemic symptoms) score and systemic reactogenicity (based on only systemic symptoms) score.

**Individual total reactogenicity score (TRC) =**

$$\left( \#of days with Grade 1 local and systemic symptoms \right)*\left( \#of different symptoms each day \right)*1+$$

$$\left( \#of days with Grade 2 local and systemic symptoms \right)*\left( \#of different symptoms each day \right)*2+$$

$$\left( \#of days with Grade 3 local and systemic symptoms \right)*\left( \#of different symptoms each day \right)*3$$

**Individual systemic reactogenicity score (SRC) =**

$$\left( \#of days with Grade 1 only systemic symptoms \right)*\left( \#of different symptoms each day \right)*1+$$

$$\left( \#of days with Grade 2 only systemic symptoms \right)*\left( \#of different symptoms each day \right)*2+$$

$$\left( \#of days with Grade 3 only systemic symptoms \right)*\left( \#of different symptoms each day \right)*3$$

Serum biomarkers

The distribution of serum biomarker response also shows consistent trends across all serum biomarkers with individuals getting dose 2 of mRNA-1647 and most of sero-positive individuals for dose 1 of mRNA-1647 showing high levels of serum biomarker response compared to other vaccine types. However, for MCP-2 the response across all vaccine types and doses is quite similar with individuals receiving dose 2 of mRNA-1647 not showing particularly higher levels compared to individuals receiving other vaccine types and dose.

Serum biomarker trajectories for all six selected serum biomarkers at multi-draws during the study period. Consistent trends are observed across all serum biomarkers with peak values occurring between 2^nd^ or 3^rd^ day and serum biomarker levels returning back to pre-vaccine levels between 4-7 days post vaccination.

**Supplementary Table 1:** Classification of solicited adverse reactions (local and systemic) by grade into a grading schema for quantifying reactogenicity

| **Reaction** | **Grade 0** | **Grade 1** | **Grade 2** | **Grade 3** |
| --- | --- | --- | --- | --- |
| Injection site pain | None | Does not interfere with activity | Interferes with activity | Prevents daily activity |
| Injection site erythema (redness) | <25 mm/  <2.5 cm | 25-50 mm/  2.5-5 cm | 51-100 mm/  5.1-10 cm | >100 mm/  >10 cm |
| Injection site swelling/  induration (hardness) | <25 mm/  <2.5 cm | 25-50 mm/  2.5-5 cm | 51-100 mm/  5.1-10 cm | >100 mm/  >10 cm |
| Axillary (underarm) swelling or tenderness ipsilateral to the side of injection | None | No interference with activity | Some interference with activity | Prevents daily activity |
| Headache | None | No interference with activity | Some interference with activity | Prevents daily activity |
| Fatigue | None | No interference with activity | Some interference with activity | Significant; prevents daily activity |
| Myalgia (muscle aches all over body) | None | No interference with activity | Some interference with activity | Significant; prevents daily activity |
| Arthralgia (joint aches in several joints) | None | No interference with activity | Some interference with activity | Significant; prevents daily activity |
| Nausea/vomiting | None | No interference with activity or 1 or 2 episodes/ 24 hours | Some interference with activity or  >2 episodes/ 24 hours | Prevents daily activity, requires outpatient intravenous hydration |
| Chills | None | No interference with activity | Some interference with activity not requiring medical intervention | Prevents daily activity and requires medical intervention |
| Fever (oral) | <38.0°C  <100.4°F | 38.0°C to 38.4°C  100.4°F to 101.1°F | 38.5°C to 38.9°C  101.2°F to 102.0°F | 39.0°C to 40.0°C  102.1°F to 104.0°F |

**Supplementary Figure 1 –** Patterns for all 63 serum biomarkers across all visits for the duration of the study for individual participants. The six selected serum biomarkers are highlighted in red box showing clear trajectories as compared to other serum biomarkers

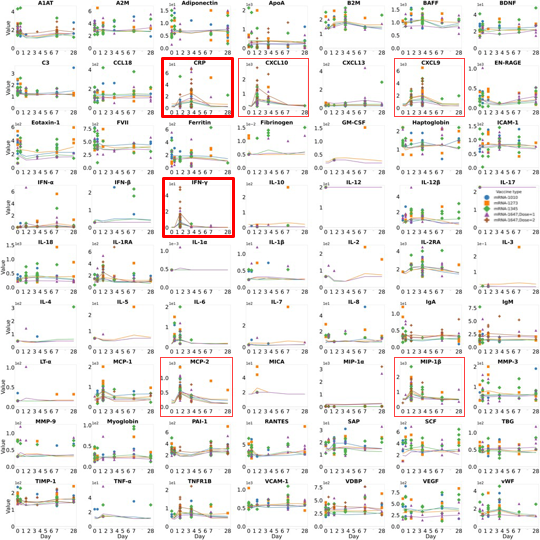

**Supplementary Figure 2 –** Patient recorded individual symptoms and respective severity across all symptoms for different vaccine types and dose

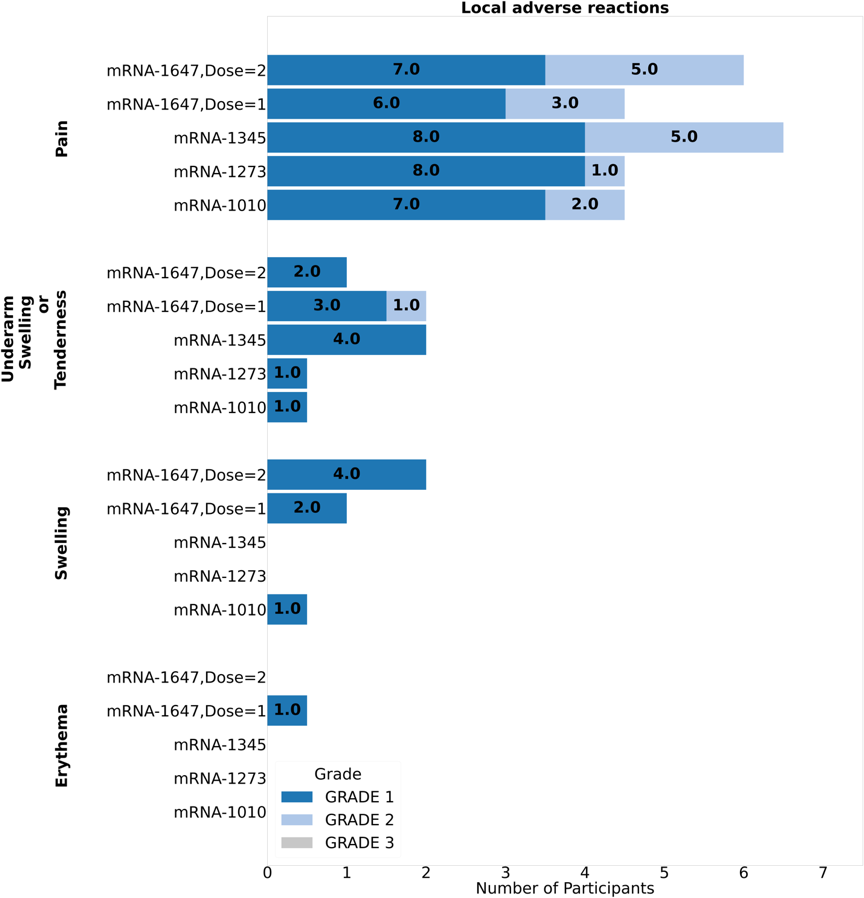

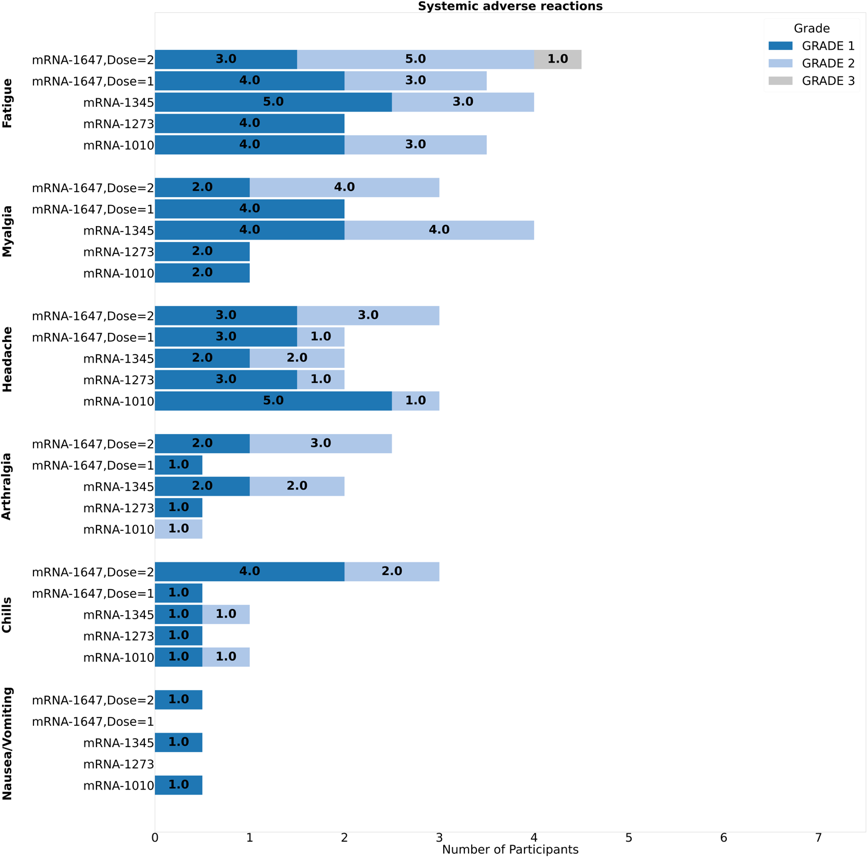

**Supplementary Figure 3 –** **(A)** Maximal change in CRP, CXCL 10, CXCL 9, IFN-$\gamma$, MCP-2, MIP-1 beta and iMCI values across vaccine types/dose. Individuals who were sero-positive for CMV at baseline are circled in red. Boxplots show the overall distribution of the response with the line representing the median value for each vaccine type and dose. Overlayed scatter plots show responses for participants who received a particular vaccine type and dose **(B)** Serum biomarker trajectories for CRP, CXCL 10, CXCL 9, IFN-$\gamma$, MCP-2, and MIP-1 beta across all vaccine types and doses over time. Solid lines represent the mean trajectories for each vaccine type whereas the markers represent the maximal change from baseline for each serum-biomarker response recorded for each individual for that vaccine type and dose.

**A)**

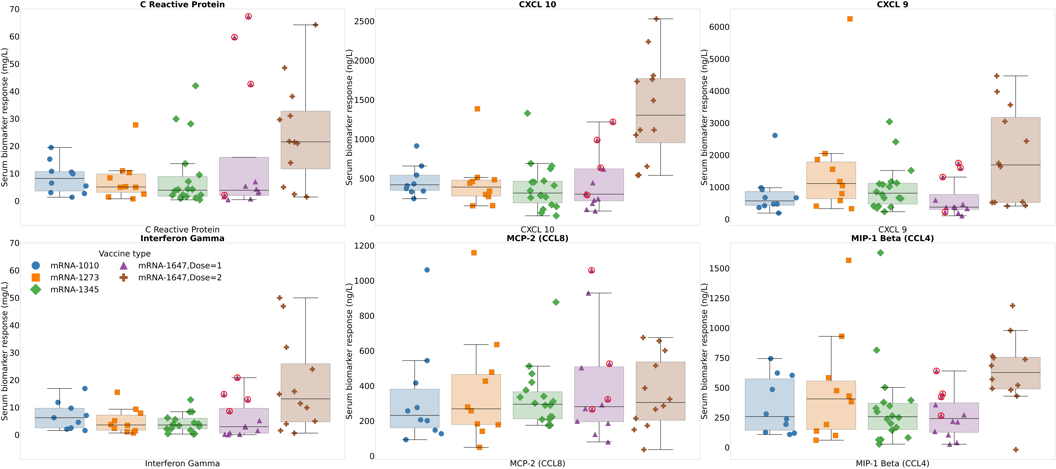

**B)**

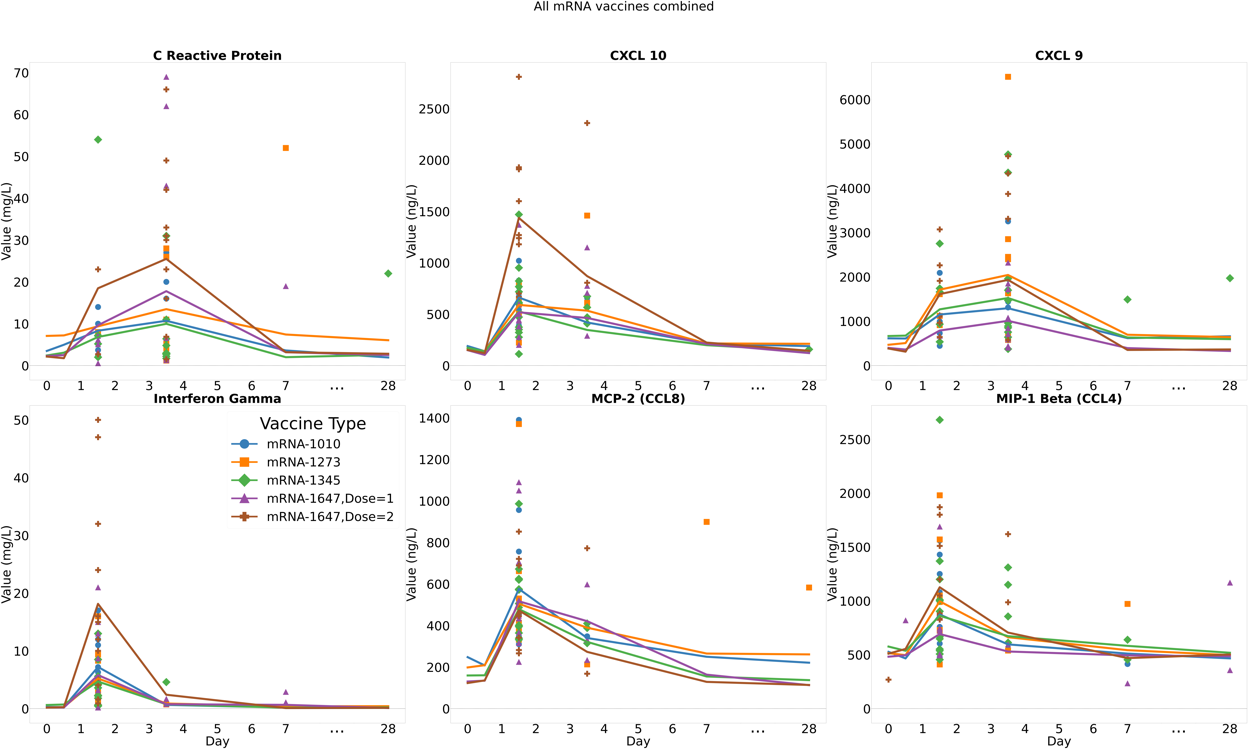

**Supplementary Figure 4 –(A)** Distribution of individual local, systemic and total reactogenicity scores across vaccine types and dose (B) Scatter plots of serum biomarkers (CRP, IFN-$\gamma$) and iMCI against local reactogenicity score and **(C)** Scatter plots of serum biomarkers (CRP, IFN-$\gamma$) and iMCI against total reactogenicity score

**A)**

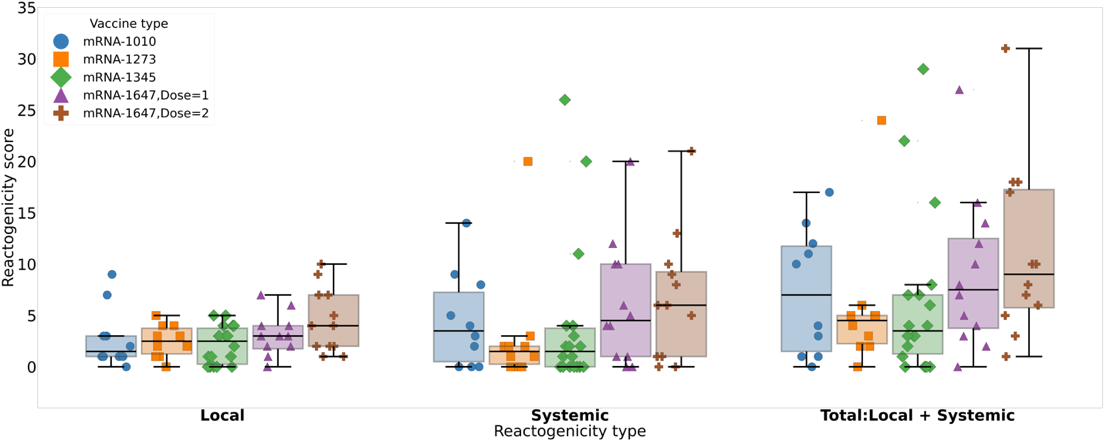

**B)**

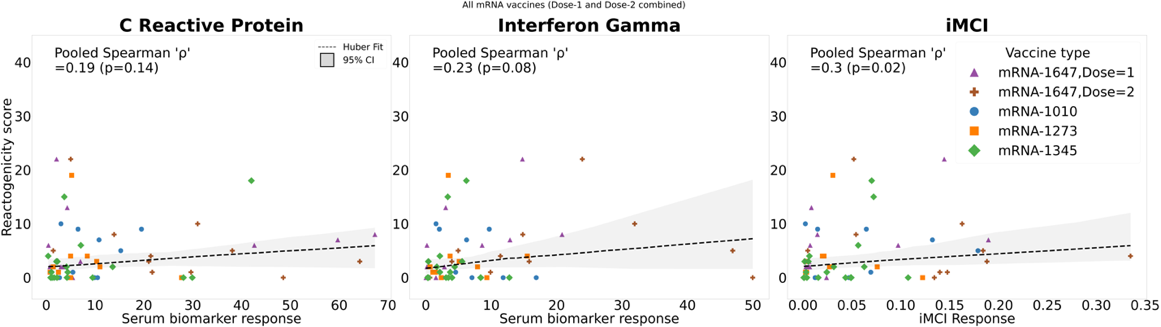

**C)**

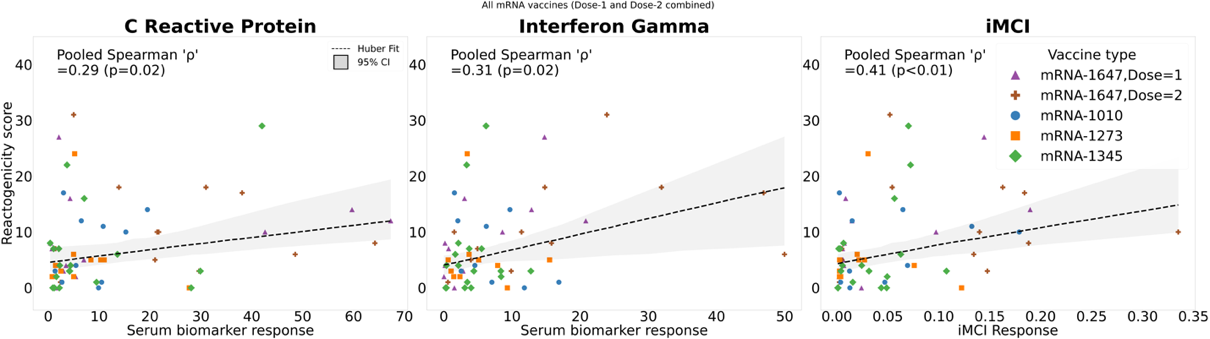
